# Drug decriminalization and population mental distress: evidence from Oregon and Washington

**DOI:** 10.64898/2026.08.10.26360079

**Authors:** Federico Allegrini, Tommaso Sonno

## Abstract

In February 2021, Oregon became the first US state to decriminalize possession of small amounts of all commonly used illicit drugs (Measure 110); twenty-four days later, Washington’s Supreme Court *Blake* ruling produced a weaker, shorter-lived decriminalization. Evaluations of these policy periods have focused on overdose deaths, with contested results; their association with the mental health of the general population is unknown. Using surveillance data on 6.3 million adult interviews (2011–2024) and synthetic control methods with permutation inference, we find frequent mental distress an estimated 2.15 percentage points higher in Oregon than in its synthetic counterfactual (largest positive gap among 45 jurisdictions; two-sided rank 2/45, *p* = 0.044, though not significant under the alternative fit-normalized statistic), with directionally consistent estimates in Washington and a joint test on the pair at *p* = 0.015. The increase concentrates in self-reported distress, among women and young adults, and is not mirrored in diagnoses, police-recorded partner violence or suicide.

## Introduction

Whether removing criminal penalties for drug possession helps or harms public health is one of the oldest questions in drug policy, and one of the least settled ^1,2^. In November 2020, Oregon voters made the United States’ first at-scale commitment to one answer: Ballot Measure 110, in force from 1 February 2021, replaced criminal penalties for possession of small, threshold-defined amounts of controlled substances (including heroin, methamphetamine and oxycodone) with a $100 citation, waivable by calling a health-assessment hotline, a condition met by roughly 1% of recipients^3,4^. The measure had a second arm: a large expansion of addiction services funded by cannabis tax revenue, whose rollout was documented as slow, with most funds reaching providers from late 2022^5,6^. What we evaluate is therefore the policy as implemented: immediate decriminalization combined with delayed treatment expansion. Twenty-four days after Measure 110 took effect, the Washington Supreme Court’s *State v. Blake* decision voided the state’s felony possession statute, producing a de facto decriminalization that the legislature converted to a misdemeanor with mandated diversion from May 2021^7,8^. Both experiments have since ended: Washington re-established criminal penalties as a gross misdemeanor from July 2023^9^, and Oregon’s House Bill 4002 restored misdemeanor penalties from 1 September 2024^10^.

A rapidly growing literature has evaluated these policies, but almost exclusively on a single margin: fatal overdoses. An early analysis attributed to Measure 110 a 23% increase in unintentional overdose deaths in 2021^11^; a subsequent synthetic-control analysis found no association with fatal overdose in either state ^12^; a later study showed that once the near-simultaneous escalation of illicit fentanyl in the Pacific Northwest’s drug supply is accounted for, no association remains ^13^; and a recent synthetic-control study estimates sustained overdose-mortality increases in both states, arguing that the standard fentanyl adjustment itself absorbs part of the policy effect ^14^. On the outcome that has dominated the public debate, the evidence remains contested.

Almost nothing is known, however, about how these policies relate to the mental health of the general population: the roughly twelve million residents of the two states, the vast majority of whom neither use criminalized drugs nor appear in any administrative dataset. A recent scoping review of drug liberalization and mental health found no study of a non-cannabis decriminalization policy and population mental health outcomes^15^; the adjacent evidence concerns cannabis legalization, with mixed findings concentrated on users, adolescents and people with psychotic disorders ^16–18^; qualitative work on Measure 110 documents changed policing and street-level dynamics but no population mental-health outcomes ^19–21^, and early evidence from British Columbia’s 2023 decriminalization concerns healthcare encounters and service access among people who use drugs, not the general population^22–24^. Prior causal work on the Oregon and Washington policies has examined overdose mortality, arrests, crime and enforcement^25^; the Oregon–Washington synthetic-control design itself has recently been applied to overdose mortality^14^. To our knowledge, this is the first study to apply it to population mental health, an outcome no prior study of any broad drug decriminalization has examined^15^; a working-paper search (NBER, SSRN, IZA, medRxiv, August 2026) found no prior application.

Population mental health is a plausible margin for such reforms to operate on, via visible drug use, perceived safety and disorder ^26^, and, for the families of people who use drugs, a population with well-documented excess health burdens^27,28^, the intensity of exposure to a relative’s use once the deterrent and the coerced “off-ramps” of the criminal justice system are removed. These channels are diffuse by construction: they require no overdose, arrest or treatment episode, and are therefore invisible to mortality records, crime reports and health-system data alike. Survey-based population surveillance is the instrument designed to capture them ^29^.

We use the largest such instrument in existence, the CDC’s Behavioral Risk Factor Surveillance System (BRFSS, 6.3 million adult interviews over 2011–2024), to track frequent mental distress, the CDC’s standard population indicator of poor mental health, in the two treated states against synthetic controls built from the other US states ^30,31^. The two adjacent experiments, one full-dose and one attenuated, allow a discipline rarely available in single-state studies: a dose–response prediction, a joint permutation test on the treated pair^32^, and a pooled design ^33^. The COVID year, deliberately excluded from all matching, provides a placebo test that any credible design must pass before 2021 gaps can be attributed to policy.

Three results follow. First, population mental distress rose in both states from 2021, in an order consistent with treatment intensity, and the increase survives adjustment, inside the counterfactual framework, for synthetic-opioid mortality (a proxy for fentanyl-era drug-market change), the pandemic and the macroeconomy. Second, it has no echo in the administrative outcomes we examine (diagnoses, police-recorded partner violence, suicide), though we quantify what each instrument could have detected. Third, the burden falls on women and young adults, with suggestive variation by household structure, not on the mostly male sanctioned population. Throughout, the estimand is the association between the 2021 policy periods as implemented and state-level population mental distress; the pattern suggests a potential population-level mental-health burden associated with these policy periods that would not be visible in overdose, arrest, diagnosis, mortality or police-recorded violence data alone.

## Results

### Design and validity

The primary outcome is frequent mental distress (FMD): the share of adults reporting 14 or more days of poor mental health in the past 30, CDC’s standard surveillance indicator ^29,34^. Secondary outcomes: mean poor-mental-health days (MHD), ever-diagnosed depression, care renounced for cost, and poor physical-health days as falsification. Over 2011–2019 Oregon’s FMD averaged 12– 13%, against a national mean near 11%. Forty-six units enter the analysis: 44 comparison units (43 states and the District of Columbia) plus the two treated states; each treated state is evaluated within its own 45-unit panel that excludes the other treated state (Methods and Supplementary S1).

For each treated state we construct a synthetic control from the 44 comparison units, matching the full pre-treatment path of the outcome over 2011–2019^30^. The COVID year 2020 is excluded from matching by design and reserved as a validity test: a synthetic control estimated without it must nonetheless track its state through the pandemic, which precedes treatment. It does: Oregon’s 2020 gap is *−*0.4 p.p., ranking 29th of 45 (FMD; MHD, 31st), and the same holds for Washington (Supplementary S2): whatever the pandemic did to population distress, it did not do measurably more of it in Oregon than in the states weighted into its synthetic control. Inference throughout is permutation-based, the standard approach for state-policy natural experiments with few treated units^12,31^: the full estimation is repeated for every state, and the treated state’s post-2021 average gap is ranked in the resulting placebo distribution. Two-sided permutation tests are primary; one-sided tests in the direction of harm, specified before the subgroup and robustness analyses were run, are a directional secondary. In other words: we build a “synthetic Oregon” as a weighted average of comparison states that reproduces Oregon’s distress history through 2019, measure how far actual Oregon departs from it after the policy, and judge that departure by rerunning the identical procedure with every comparison state in Oregon’s place. Because significance can depend on how the departure is scored, we score it two ways throughout and report both. We avoid a conventional two-way fixed-effects event study as the primary design because Oregon fluctuated 1–2 p.p. above the national mean already in 2012–2018, so pre-period coefficients against any base year inherit that fluctuation; the synthetic control instead builds the counterfactual from the full pre-path, and its pre-trend test, the gap staying inside the placebo band through 2020, is met (Figs. 1 and 2).

**Figure 1:**
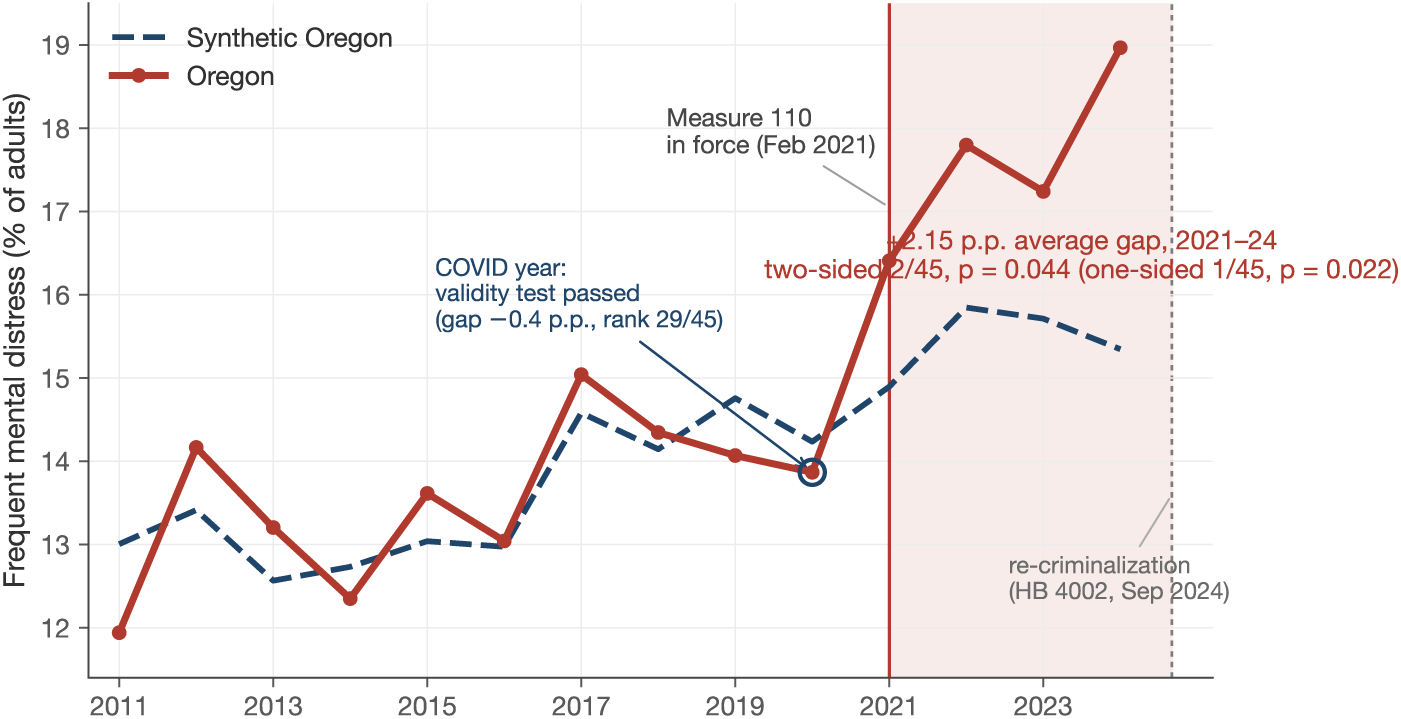
Oregon versus synthetic Oregon: frequent mental distress, 2011–2024.

**Figure 2:**
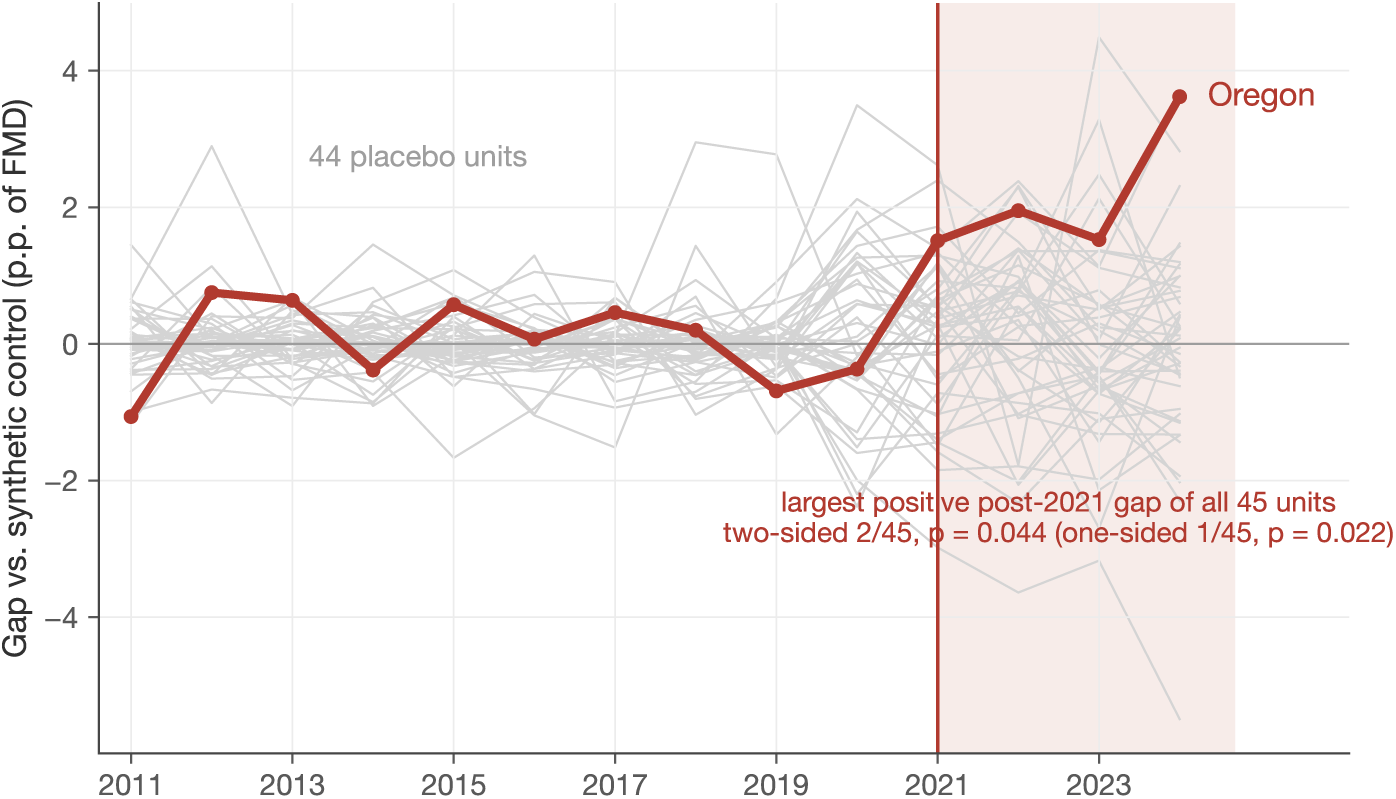
Oregon’s gap against the placebo distribution. Notes: Treated–synthetic gap for Oregon (dark) and each comparison state (grey). Oregon’s average post-2021 gap (+2.15 p.p.) is the largest positive gap of all 45 units (two-sided *p* = 0.044; one-sided *p* = 0.022); its 2020 gap sits at the center of the distribution. Under the alternative RMSPE-ratio statistic Oregon ranks 11/43 (*p* = 0.256); see Table 1.

**Table 1:**
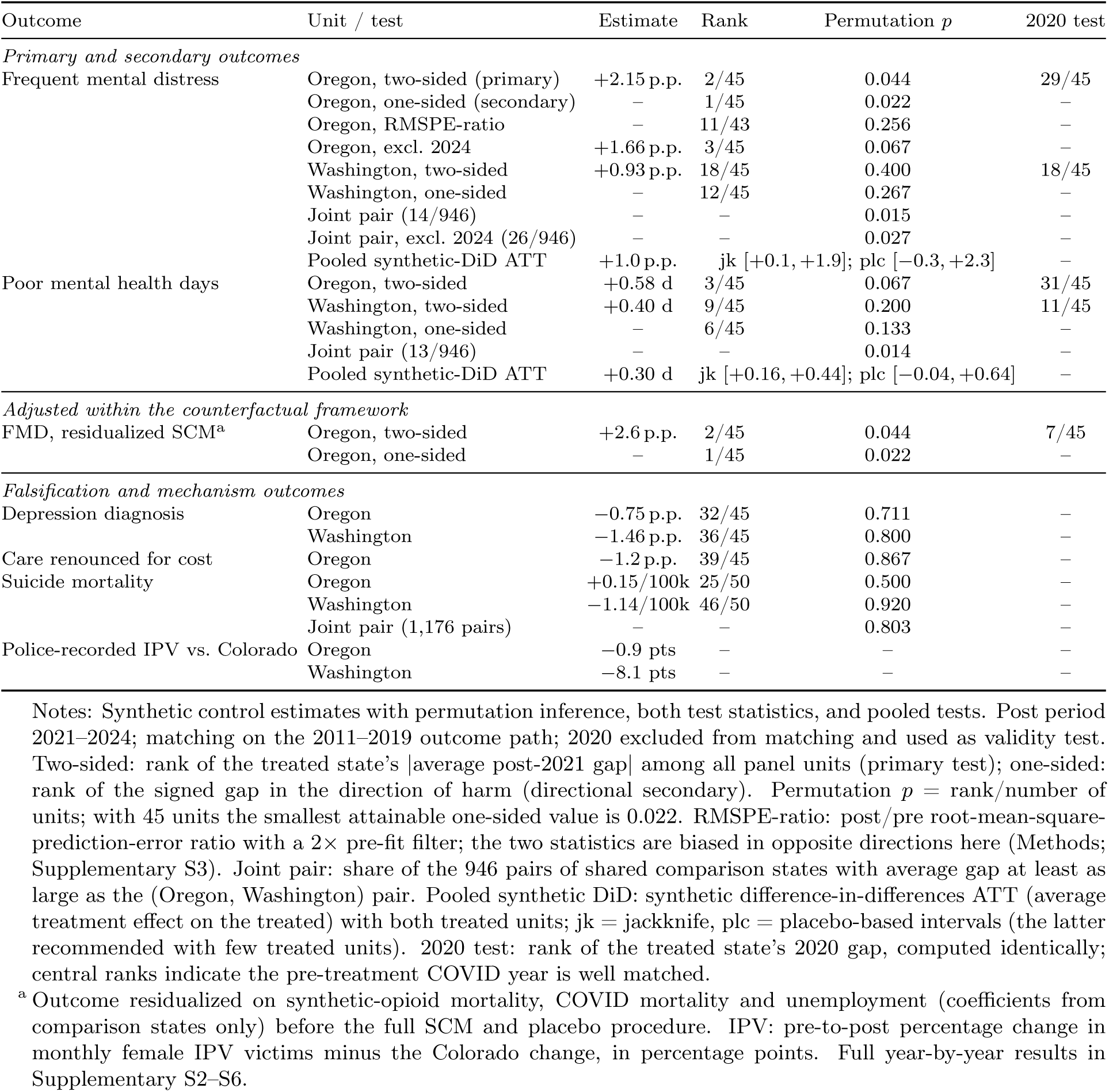
Main results.

### Distress rises from 2021 in both states

From 2021 Oregon’s FMD gap turns positive and stays positive for four consecutive years (+1.5, +2.0, +1.5, +3.6 p.p.; annual placebo ranks 4, 4, 5 and 2 of 45). On the statistic that rewards persistence rather than single-year spikes, the average post-treatment gap, Oregon has the largest positive gap of all 45 units: +2.15 p.p., roughly 14% of the counterfactual level; the two-sided permutation *p* is 2*/*45 = 0.044 (one-sided, in the direction of harm: 1*/*45 = 0.022, the smallest value the 45-unit design can produce) (Figs. 1 and 2, Table 1; raw series for all 46 units in Supplementary Fig. S1). These are rank statements, not large-sample *p*-values: the summary is that Oregon is a strong state-level anomaly aligned with policy timing. In absolute terms this is on the order of 70,000 additional Oregon adults above the frequent-distress threshold in an average post-reform year. Poor-mental-health days, a transformation of the same survey item rather than an independent outcome, rise by +0.58 days (12% of the counterfactual; two-sided 3*/*45 = 0.067, one-sided 2*/*45 = 0.044). The estimate is not driven by any single year: leaving out each post year in turn, Oregon ranks 2/45, 2/45, 2/45 and 3/45. Under the alternative RMSPE-ratio statistic, which normalizes each state’s post gap by its pre-period fit (the conventional 2*×* fit filter excludes two ill-fitting units, leaving 43), Oregon ranks 11 of 43 (*p* = 0.256): with two treated units, significance depends partly on the choice of test statistic. The two statistics are biased in opposite directions here, because 31 of 44 placebo runs fit their state better than Oregon’s (Supplementary S3 reports the diagnostic and both statistics in full); we report both and do not rest the paper’s claim on either alone. Correcting for testing across the three-outcome family (max-*t* permutation), the result stands at rank 3/45 (*p* = 0.067); excluding 2024, at rank 3/45 (*p* = 0.067).

Washington provides a weaker-policy comparison whose point estimates are directionally consistent with Oregon’s. Its treatment was weaker on both margins (judicially imposed rather than voted, and converted to a misdemeanor with mandated diversion after eleven weeks^8^), and its estimated gap is about half of Oregon’s (+0.93 p.p. FMD; MHD +0.40, rank 9/45), positive in every post year, extreme in none. Individually, Washington is not distinguishable from the placebo distribution (*p* = 0.40 for FMD, two-sided); a smaller effect is less likely to appear extreme in a noisy 45-unit placebo distribution, so Washington is best read through its sign, its timing, and its contribution to the joint analyses, not as a stand-alone test. The probability that a randomly chosen pair of the 44 shared comparison units matches or exceeds the treated pair’s joint average gap is 14/946 (*p* = 0.015) for FMD and 13/946 (*p* = 0.014) for MHD ^32^; excluding 2024, 26/946 (*p* = 0.027). Pooling the two treated units in a synthetic difference-in-differences design (a version of the method that averages the two treated states and delivers confidence intervals) ^33^ gives an average effect of +1.0 p.p. for FMD, with a 95% interval of [+0.1, +1.9] under jackknife standard errors and [*−*0.3, +2.3] under the placebo-based procedure recommended with few treated units, which includes zero; for MHD, +0.30 days, [+0.16, +0.44] and [*−*0.04, +0.64] respectively. The pooled estimates sit below Oregon’s because they average the full-dose treatment with Washington’s attenuated one, and the joint test assumes both units share the treatment mechanism, an assumption revisited in the Discussion (Fig. 3). The treated pair also shows pre-period deviations, most notably 2013 (*p* = 0.039 FMD; *p* = 0.001 MHD), which we report here because bands this tight have power against pre-period noise too (Supplementary S2).

**Figure 3:**
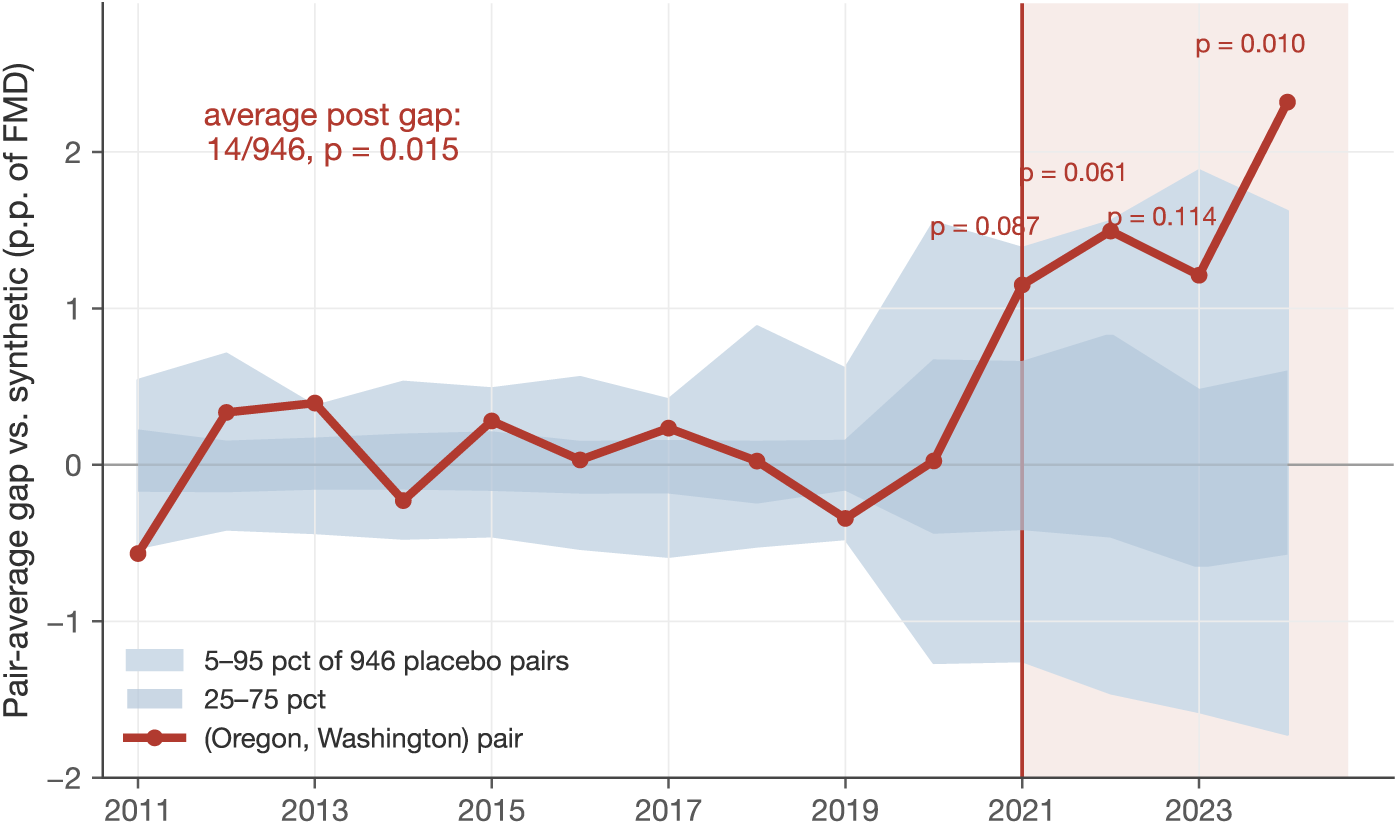
The treated pair against all 946 placebo pairs. Notes: Average FMD gap of (Oregon, Washington) by year versus the 5–95 percentile band of all pairs of shared comparison states. Annual *p*: 0.087, 0.061, 0.114, 0.010 (2021–24); average post gap, *p* = 0.015; excluding 2024, *p* = 0.027.

### More distress, not more diagnoses, not more suicide

Depression diagnoses do not rise in Oregon (gap *−*0.75 p.p., rank 32/45; Fig. 4); in Washington the diagnosis series drifts negative over the post period (*−*1.46 p.p., rank 36/45). Two qualifications discipline this null: the BRFSS item is a lifetime-diagnosis stock (even a 20% rise in new diagnoses would move it by about one percentage point at most over four years, near the detection floor; Supplementary S2), and this outcome has the weakest pre-period fit of the three. What the contrast supports: substantially more reported suffering without a proportional trace in diagnoses, while care renounced for cost does not rise (rank 39/45), so affordability is not the missing link. Suicide mortality, from complete death certificates with the identical design, shows no effect: Oregon rank 25/50 (*p* = 0.50), Washington 46/50, joint *p* = 0.80; given Oregon’s suicide counts (883 deaths in 2022, a crude rate of 20.8 per 100,000), gaps smaller than roughly 7% of the baseline rate would be undetectable at conventional ranks (calculation in Supplementary S6), so this null excludes large effects, not all effects. This null does not conflict with the overdose-mortality increases estimated for these states^14^, which count drug poisonings of all intents, dominated by unintentional deaths and only partially overlapping the suicide codes analyzed here. The rise is in moderate, sustained distress that does not surface in diagnoses, death certificates or police records.

**Figure 4:**
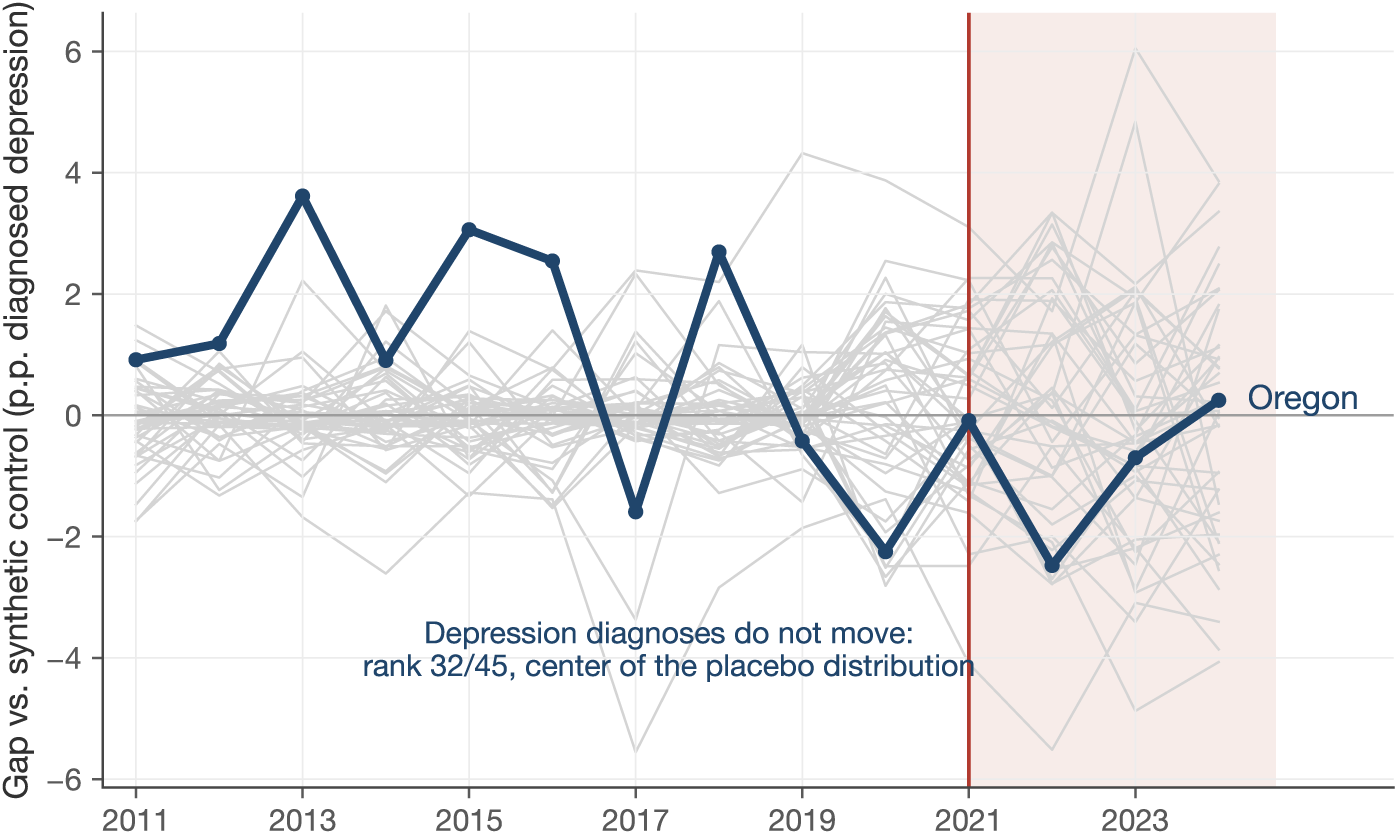
No comparable movement in depression diagnoses. Notes: Identical design, outcome = ever-diagnosed depression; Oregon gap *−*0.75 p.p., rank 32/45. This outcome is a lifetime-diagnosis stock with limited sensitivity to flow changes, and has the weakest pre-period fit of the three outcomes (Supplementary S2).

### Who reports the increase: exploratory heterogeneity

Oregon’s administrative records show who the policy directly touched: citations replaced roughly 4,100 annual criminal convictions, arrests for possession fell sharply, and about three-quarters of the cited were men (Supplementary S7). The distress increase, in contrast, is clearest among women: on the female-only panel Oregon ranks 1/45 on poor-mental-health days (*p* = 0.022) and 2/45 on FMD, while the aggregate male panel is unremarkable (ranks 13–14/45). By age, the young bear the brunt (18–34: rank 4/45; over-65: null), with a monotone age gradient in the microdata (+0.32 days for ages 18–34 versus +0.04 for 65+).

Household structure adds texture that we label exploratory, quantifying both sides of the label: heterogeneity is formally present (microdata omnibus test across the four sex-by-partnership groups, *F* = 26.9, *p <* 0.001, state-clustered), but no single subgroup survives family-wise correction (max-statistic across the eight household panels: 9/45, *p* = 0.20; Supplementary S4). The two groups that rank high are partnered women (3/45, *p* = 0.067) and single men (4/45, *p* = 0.089), the latter being the group that contains most people who use criminalized drugs; partnered men (22/45) and single women (12–17/45) are unremarkable. Among partnered women aged 18–54, the gap is largest for those without children at home (+4.4 p.p., rank 3/45) and absent for mothers. Separation and divorce rates do not move, so the pattern is not a family-composition artifact. We read this configuration as suggestive of a household-exposure channel; establishing it would require household-level exposure data that BRFSS does not contain.

A physical-violence mechanism finds no support in police records. We construct monthly police-recorded intimate-partner-violence (IPV) series from FBI incident-level crime records (female victims of assault by a partner or ex-partner) for Oregon, for Washington, and for Colorado as an untreated comparison, chosen because it shares the treated states’ legal-cannabis environment and urban profile, never decriminalized other drugs, and reported complete incident-based crime data throughout 2018–2024 (Supplementary S5). Police-recorded IPV is flat through both treatments: pre-to-post changes are +6.3% in Oregon, *−*0.9% in Washington and +7.2% in the Colorado comparison (*−*0.9 and *−*8.1 points relative to Colorado, respectively), with no discontinuity at the treatment dates, no reversal after re-criminalization, and a broadly stable IPV share of all female assault victimizations. The result is not an artifact of agency composition: restricting to the panel of agencies reporting in every year 2018–2024 (which covers 97–99% of recorded IPV victims in all three states) leaves the changes essentially unchanged (+4.7%, *−*1.7%, +6.8%; Supplementary S5). Two caveats bound this null: recorded IPV is conditional on reporting to police, which the policy environment could itself affect; and Portland’s domestic-violence 911 dispatches double over the period, a 2020-onset rise best read against flat statewide victim records as a dispatch phenomenon (Supplementary S5). These records show no detectable increase in partner violence; they do not rule out changes in unreported violence or other household stressors.

### Robustness checks for alternative explanations

One distinction organizes this section: measurable confounders (fentanyl mortality, COVID mortality, unemployment, survey composition, geography) can be adjusted for or tested; unmeasured features of the policy environment itself (homelessness, visible public use, service strain) are plausibly downstream of the policy, and adjusting for them would remove part of what is under study. The checks below address the first group.

#### Fentanyl, COVID and the macroeconomy, inside the counterfactual framework

Illicit fentanyl reached Oregon essentially together with the policy (synthetic-opioid deaths rose from 2.0 to 31.5 per 100,000 between 2019 and 2023), and adjusting for it eliminated the overdose findings^13^. We therefore partial synthetic-opioid mortality, COVID mortality and unemployment out of the outcome, with coefficients estimated on comparison states only, and re-run the entire synthetic-control and placebo machinery on the residuals: Oregon remains the most extreme unit (rank 1/45 one-sided, 2/45 two-sided; residualized gap +2.6 p.p.), although the 2020 placebo on the residualized outcome is less central (rank 7/45), reflecting the noise that 2020 unemployment injects. A distinct pandemic rival gets its own test: Oregon and Washington stayed relatively restrictive into 2021–22, and prolonged closures could mimic our signature. Adding average policy stringency and school-closure intensity (OxCGRT indices) to the adjustment set changes nothing (Oregon rank 1/45 one-sided, 2/45 two-sided; gap +2.6 p.p.). And the school-closure account predicts the burden should fall on parents; the opposite holds: the largest subgroup gap is among partnered women *without* children (+4.4 p.p.), mothers show none (+0.3; Supplementary S4). A ridge-augmented synthetic control^35^, which corrects for residual pre-period imbalance, returns the same answer (gap +2.2 p.p., rank 1/45 one-sided, 2/45 two-sided), while moving economic covariates into the predictor set worsens Oregon’s pre-fit and attenuates the gap (+1.5 p.p., rank 3/45 one-sided, 7/45 two-sided; Supplementary S3). A two-way fixed-effects specification with the same controls, used only as a coefficient-stability check given its unsuitable pre-trends here, attenuates post-2021 coefficients by 2–31% without changing their profile (Fig. 5; FMD 2024: +3.1 to +2.7 p.p.).

**Figure 5:**
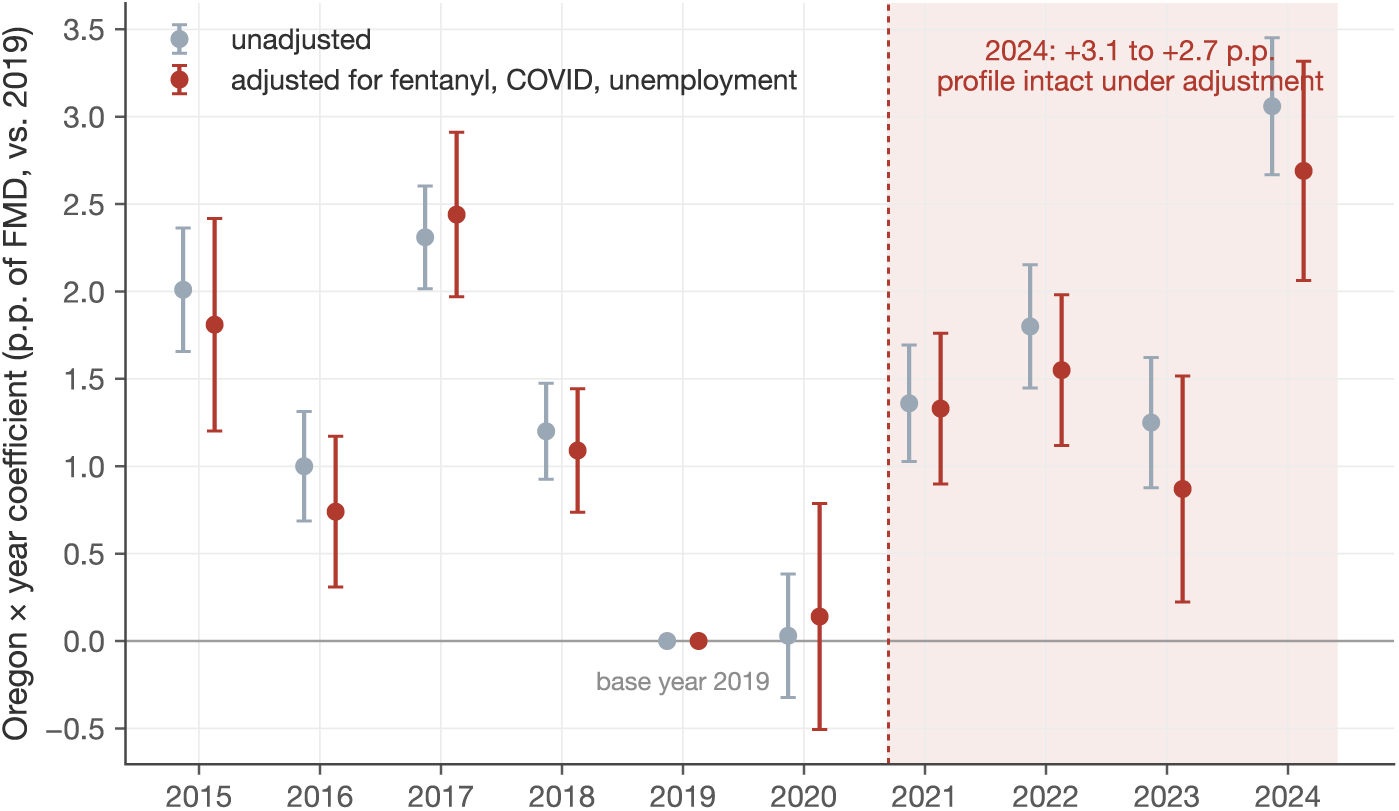
Coefficient-stability check under confounder adjustment. Notes: Two-way fixed-effects coefficients on Oregon*×*year with and without controls for synthetic-opioid mortality, COVID mortality and unemployment. The non-flat pre-2021 coefficients illustrate why this design is unsuitable as the primary specification here (Oregon fluctuates 1–2 p.p. above the national mean relative to the 2019 base year); the figure’s only role is to show that adjustment changes post-2021 coefficients by 2–31% without altering their profile. The primary covariate-adjusted evidence is the residualized synthetic control in Table 1.

#### COVID

Beyond the 2020 validity test, including 2020 among the matching predictors, the least favorable choice for a pandemic story, strengthens the result (rank 1/45; Supplementary S2).

#### Survey artifacts

A synthetic control on the log number of BRFSS respondents puts Oregon at rank 14/45: no differential sampling. Re-weighting Oregon’s post-2020 respondents to their pre-2020 demographic composition (entropy balancing on sex, age, race, education, income) leaves the increase intact. American Association for Public Opinion Research (AAPOR) combined response rates for both treated states track the national median throughout the post period, with deviations well inside their pre-treatment ranges (Supplementary S8). The physical-health falsification outcome, run through the identical placebo machinery, is null: Oregon ranks 21/45 on poor physical-health days (*p* = 0.47; frequent physical distress, 30/45), with a modest upward drift confined to 2023–24 (+0.19 and +0.52 days) that never approaches the placebo extremes. An independent federal instrument points the same way as the primary outcome: in the Census Household Pulse Survey, Oregon ranks at or near the top ten jurisdictions (6th–11th of 51) on anxiety–depressive symptoms in every year 2021–2024; because that survey only begins in April 2020, it can corroborate Oregon’s elevated level but not the pre–post change (Supplementary S8).

#### Geography within Oregon

If the divergence reflected Portland-specific urban conditions (post-2020 disorder, homelessness, a visible fentanyl scene) rather than a statewide policy environment, it should be absent outside the Portland metro; it is not. In the CDC’s metropolitan SMART subsample (Selected Metropolitan/Micropolitan Area Risk Trends; Supplementary S3), FMD rises from 2017–19 to 2021–24 by +4.1 p.p. in the Portland metro area and by +4.7 p.p. in Salem; combined with the statewide series, these imply an increase of roughly +2 p.p. in the remainder of the state. The available metropolitan estimates thus show increases in both Portland and Salem and imply a smaller increase elsewhere in Oregon, which argues against an exclusively Portland-specific account, although these sub-state comparisons are descriptive and the Portland-area estimate includes Clark County, Washington (Supplementary S3).

#### Neighbors

Idaho and California, adjacent and never decriminalizing, are unremarkable (ranks 30 and 17 of 45): there is no generic West Coast shock. Nevada appears extreme (rank 3/45) but with the wrong signature: its gap opens in 2020, before any treatment, alongside the country’s worst pandemic unemployment shock (13.5%), and it fails the 2020 validity test that Oregon passes.

#### Specification

Recomputing the entire placebo distribution under pre-specified variants (three-year predictor averages, shortened pre-period, 2020 included) keeps Oregon between rank 1 and 4 of 45 and the joint test between *p* = 0.001 and 0.058^36^. A summary mental-health index ranks 2/45; the joint test restricted to same-census-division pairs gives *p* = 0.018–0.035. On a quarterly panel, Oregon ranks 2/45 over the quarterly post period, and the first half of 2024, entirely under Measure 110, already shows a +2.7 p.p. gap, so the strong 2024 is not an artifact of the post-reversal months (Supplementary S3).

## Discussion

Oregon’s decriminalization period was associated with a persistently higher level of population mental distress relative to its synthetic counterfactual, with an onset that coincides with the policy: roughly one additional adult in fifty above the frequent-distress threshold (about 70,000 adults on a base of roughly 3.3 million), while depression diagnoses, police-recorded partner violence and suicide mortality show no comparable movement. The strength of this evidence is the conjunction: onset at treatment in both states, a demanding 2020 placebo passed, survival under covariate adjustment inside the counterfactual framework, stability across pre-specified variants. Its weakness is equally identifiable: with two treated units, significance depends partly on the test statistic, and we report the less favorable choices alongside the favorable ones.

The configuration of results carries information about mechanism, which we read cautiously. The population the policy directly touched (those cited, three-quarters male) shows no clear distress increase in the aggregate male panel, although single men rank high among subgroups. The clearest movements are among women and young adults, with exploratory heterogeneity suggesting a role for household structure. What increased is the layer of population suffering that only surveillance surveys measure: no additional police-recorded violence, no additional suicides, no additional diagnoses. This is why an evaluation literature focused on overdose mortality, understandably given the fentanyl crisis, could not have detected it^11–13^. We treat the “psychological burden of proximity” reading as a hypothesis consistent with, but not identified by, the available evidence: the clinical literature on families affected by substance use^27,28,37^ and the literature linking perceived neighborhood disorder to anxiety and depression ^26,38^ make it plausible, the sex-age-household pattern is compatible with it, and Supplementary S4 sets out, channel by channel, the evidence for it, the evidence against it, and what each available measure can and cannot capture.

Washington’s own re-criminalization (July 2023) ^9^ offers an in-sample check, and we report its result: Washington’s gap does not visibly revert in 2023–24 (+0.90, +1.02 p.p.). Three readings are possible: the dose was already low after May 2021; annual data cannot resolve a mid-2023 change; or the Washington gap was never policy-driven, shifting evidentiary weight onto Oregon. Current data cannot distinguish these.

The findings imply that evaluations of decriminalization should include population mental-health surveillance alongside overdose, enforcement, treatment and public-safety outcomes. On overdose mortality the evidence is now contested, with credible estimates on both sides of the null ^12–14^; the policy also delivered real, documented benefits, removing roughly four thousand criminal convictions per year, with the largest proportional reductions projected for Black and Indigenous Oregonians in the state’s racial-impact analysis^39^. Its critics correctly note that public drug use became more visible. The margin that moved in our data, diffuse psychological burden in the general population, was not the subject of either side’s argument. For the European debates, where the Portuguese model remains the touchstone ^1^, we caution against direct read-across in either direction: Oregon’s implementation, drug supply and services context differ from Portugal’s in ways that plausibly matter. The most generalizable implication is procedural: evaluations of future reforms should measure population mental health prospectively, even where the expected sign and magnitude are uncertain.

Several limitations follow. With two treated units, all inference is permutation-based; ranks among 45 units have irreducible granularity, and the field-standard alternative statistic is non-significant for Oregon alone. Self-reported distress has reporting margins. Secular shifts in the willingness to label distress as a problem (concept creep^40^) are national in scope and difference out by construction; the rival we cannot fully exclude is state-specific: the policy itself reframed drug use as a health issue and dominated local media, which could alter disclosure independently of experienced distress. The survey-composition, sampling and falsification tests bound this concern, and a first triangulation is now in place: in the Census Household Pulse Survey, an independent instrument with a different mode and sampling frame, Oregon ranks at or near the top ten jurisdictions (6th–11th of 51) on anxiety–depressive symptoms in every year 2021–2024, which is difficult to reconcile with a BRFSS-specific reporting artifact; because that survey begins only in April 2020, however, it cannot verify the pre–post change itself (Supplementary S8). A second, drug-native triangulation points the same way: the federal substance-use survey (NSDUH) measures drug use and mental illness in the same instrument, and there Oregon ranks second or third nationally on past-month illicit-drug use other than marijuana in every wave, and rises from third to first on any mental illness between its 2021–22 and 2022–23 waves (Supplementary S8). The two-way fixed-effects stability check suggests a modest share (2–31%) of the post-2021 coefficients travels with the drug-supply and macroeconomic context, while the residualized synthetic-control gap is, if anything, slightly larger (+2.6 p.p.). Finally, 2024, the strongest year, is also the year the physical-health falsification outcome drifts, and the estimate excluding 2024 is weaker (rank 3/45, *p* = 0.067), although the joint test without 2024 remains unlikely under the null (*p* = 0.027). Last, the design compares states; a follow-up exploiting within-state variation in exposure (county citation intensity, the staggered rollout of funded treatment sites) would relax the assumption that nothing else Oregon-specific changed in 2021, and requires sub-state mental-health data that public BRFSS files do not provide.

Oregon re-criminalized possession from September 2024^10^. If the mechanism is real, Oregon’s distress gap should begin to shrink when the 2025 surveillance data are released (expected August– September 2026). The prediction, primary statistic and decision rule for this reversal test are specified in Supplementary Section S9 and deposited in a public registry before the data exist (OSF; registration identifier in Supplementary S9). The pre-registered test will either strengthen the chain of evidence (onset with the policy, dose ordering across states, reversal with repeal) or weaken it, on the record either way; both outcomes are informative, and both are on the record in advance.

## Methods

### Data and outcomes

We use BRFSS public-use microdata, 2011–2024 (*N* = 6,347,000 adult interviews; CDC final weights _LLCPWT throughout) ^41^. The primary outcome, frequent mental distress, is **1**[MENTHLTH *≥* 14], where MENTHLTH is days of poor mental health in the past 30 (88 recoded to 0; 77/99 to missing): the CDC surveillance standard, with documented reliability and validity ^29,34,42,43^. Secondary outcomes: MHD (mean MENTHLTH days); depression ever diagnosed (ADDEPEV, harmonized across waves); poor physical-health days (PHYSHLTH, same recodes) as falsification; care renounced for cost (MEDCOST). State-year cells aggregate all respondents with survey weights; subgroup panels (sex, age bands, marital status *×* sex, children in household) are built from the same extraction with identical recodes (Supplementary S1). Forty-six units enter the analysis: 43 comparison states plus the District of Columbia (44 comparison units in total), plus Oregon and Washington; the five states with incomplete outcome series (Florida, Kentucky, New Jersey, Pennsylvania, Tennessee) are dropped identically for treated and placebo estimations, and each treated state is evaluated within its own 45-unit panel that excludes the other treated state. Item non-response on MENTHLTH is below 2% in all years and is dropped item-wise; no imputation is performed. The 2024 public-use file was released with modifications, and Tennessee was unable to meet inclusion requirements; Tennessee is among the five states our balanced-panel rule already excludes, and all variables used here (MENTHLTH, PHYSHLTH, depression and care-cost items, sex, marital status, age bands, children, weights) are present and populated in the 2024 file. One rival we do not test directly is wildfire-smoke exposure, which is Oregon-specific in some years; we flag it as untested rather than adjusted for. Two features of the data nonetheless cut against a smoke account: the most severe smoke exposure of the sample period (September 2020) falls in the excluded validity year, in which Oregon’s distress gap is *−*0.4 p.p. and centrally ranked, while the post-2021 gaps arise in years with milder smoke seasons and are largest in 2024; and the demographic signature (concentration among partnered women without children and young adults rather than older adults) does not match the population most affected by smoke-related health burdens.

### Treatment definitions

Oregon is treated from survey year 2021 (Measure 110 effective 1 February 2021) ^3^; Washington from 2021 (*State v. Blake*, 25 February 2021) ^7^, converted to a misdemeanor with mandated diversion from 13 May 2021 (ESB 5476) ^8^ and to a gross misdemeanor from July 2023 (2E2SSB 5536) ^9^; we therefore treat Washington as a second, lower-dose unit throughout, and exclude each treated state from the other’s comparison pool. The post period is 2021–2024. Oregon’s HB 4002, signed 1 April 2024, restored misdemeanor penalties from 1 September 2024 ^10^; 2024 is thus 8/12 treated months, and a quarterly design isolates 2024H1 (Supplementary S3).

### Synthetic control

For outcome *y* and treated state *s*, comparison-state weights (the “donor pool” of the synthetic-control literature) minimize the distance between treated and weighted-comparison outcomes on predictors *y*(2011)*, …, y*(2019): the full pre-treatment path, with 2020 excluded by design ^30,31^. The estimand is the average treatment effect on the treated (ATT), estimated as the average post gap 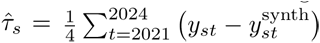. Comparison-state weight vectors and pre-period fit diagnostics for both treated states are reported in Supplementary S3.

### Policy endogeneity

Adoption was not random: the electorate that passed Measure 110 plausibly differs in drug-market conditions and attitudes toward drug policy. Selection on the pre-treatment level and trend of the outcome is absorbed by construction, because the counterfactual is matched on Oregon’s 2011–2019 outcome path; the residual threat is adoption timed to coincide with state-specific future shocks, which the out-of-sample 2020 test probes but cannot eliminate.

### Permutation inference

The identical procedure is repeated for every state in the panel; the permutation *p*-value is the treated state’s rank of *τ̂* divided by the number of units (45). Two-sided rank tests (ranks of *|τ̂|*) are primary; one-sided tests in the direction of harm, the theoretically motivated direction, are reported as a secondary; the direction was specified before the subgroup and robustness analyses were run but is not covered by a dated registry (the pre-registered protocol in Supplementary S9 concerns the reversal test), which is one reason the two-sided test is primary. We report both the gap-rank statistic and the RMSPE-ratio statistic, which are biased in opposite directions when the treated unit’s pre-fit differs from the placebo median (Supplementary S3): 31 of 44 placebo runs fit better than Oregon’s (pre-RMSPE 0.0059 vs. placebo median 0.0043), which mechanically inflates well-fitting placebos’ ratios and penalizes Oregon under the ratio statistic, while a worse-fitting treated unit can generate larger post gaps under the gap statistic. The 2020 validity rank is computed identically on the 2020 gap. The joint test compares 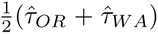 with the same statistic for all 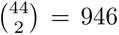 pairs of the shared comparison units ^32^. Leave-one-year-out estimates recompute *τ̂* and its rank excluding each post year in turn. Specification variants (three-year predictor averages, shortened pre-period, 2020 included among predictors), fixed before the subgroup and mechanism analyses were run, recompute the entire placebo distribution under each variant^36^; the averaged-predictor variants also answer the known objection to all-lags predictor sets ^44^. A further variant moves genuine covariates into the predictor set: averaged outcome lags (2011–2015, 2016– 2019, plus the 2019 value) together with pre-period unemployment, median household income and poverty rates, again recomputing the full placebo distribution (Supplementary S3).

### Covariate adjustment within the counterfactual framework

We regress the state-year outcome on synthetic-opioid mortality (CDC VSRR, T40.4), COVID mortality, unemployment (BLS LAUS) and, in an extended specification, average COVID policy stringency and school-closure intensity (OxCGRT state-level indices, annual means 2020–2022), with year effects, using comparison states only; structural zeros are assigned where the series did not exist (synthetic-opioid deaths before 2015, COVID-era variables outside 2020–2022). We then subtract the covariate component from all states and re-run the full synthetic-control and placebo procedure on the residualized outcome.

### Ridge-augmented synthetic control

We follow Ben-Michael, Feller and Rothstein ^35^ and reestimate the counterfactual with a ridge-augmented synthetic control, which corrects the standard prediction for remaining pre-period imbalance using a ridge regression of comparison-unit post-period outcomes on their pre-period histories, with the penalty chosen by leave-one-out cross-validation among comparison units, and we recompute the full 45-unit placebo distribution under the augmented estimator.

### Conformal inference

We implemented the conformal procedure of Chernozhukov, Wüthrich and Zhu ^45^, which refits the counterfactual under the null on all periods and applies a moving-block permutation to the residuals. We report the conformal diagnostics rather than use the conformal *p*-value as a decision statistic, because in this short-post-window setting (four post years in a thirteen-year series, with 44 comparison units) the null refit absorbs the post-2021 divergence into the weights, post-period residuals shrink from +1.6/+3.7 p.p. to +0.2/+1.1 p.p., at the cost of a 28% worse pre-period fit (implementation-specific fit; the primary design’s pre-RMSPE is 0.59 p.p.). Under that refit the procedure loses its ability to discriminate between the null and the estimated divergence ^45^. Diagnostics in Supplementary S3.

### Pooled synthetic difference-in-differences

sdid^33^ on the panel including both treated units; we report both jackknife and placebo-based standard errors, and note that the placebo-based procedure is the recommended one with very few treated units, consistent with placebo-based inference for settings with a small number of policy changes ^46^; its validity rests on treating the treated units’ shocks as exchangeable with (homoskedastic relative to) the comparison units’ shocks.

### Two-way fixed-effects stability check

A regression of state-year outcomes on Oregon*×*year indicators with state and year fixed effects, population-weighted, SEs clustered by state, with and without the three covariates above. Given Oregon’s non-flat pre-period relative to any single base year, this design serves only to check coefficient stability under confounder adjustment; the synthetic control is the primary design.

### Mechanism data

#### Enforcement

Oregon Judicial Department Measure 110 statistics and Criminal Justice Commission reports (citations, convictions replaced, arrests, appearance rates, demographic composition; sources and series in Supplementary S7) ^4,39^. *Intimate-partner violence*: FBI National Incident-Based Reporting System incident-level bulk files for Oregon, Washington and Colorado, 2018– 2024; IPV defined as aggravated or simple assault (offense codes 13A/13B) against individual female victims whose victim–offender relationship is spouse, common-law spouse, ex-spouse, boyfriend/girlfriend or ex-relationship; monthly victim counts; the parser exactly reproduces an independent earlier extraction (Supplementary S5). All three states reported incident-based data throughout 2018–2024, predating the FBI’s 2021 collection transition; agency-composition checks are reported in Supplementary S5. *Suicide*: CDC WONDER underlying-cause mortality (Injury Intent = Suicide), 51 jurisdictions (50 states and DC), 1999–2024 final plus 2025 provisional; each treated state is evaluated within a 50-unit panel excluding the other, and the joint test uses the 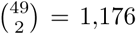 pairs of the 49 shared jurisdictions (Supplementary S6). *Municipal dispatch*: Portland Police Bureau open dispatch data (Supplementary S5).

### Robustness battery

Sex- and age-specific panels; marital *×* sex and children *×* partnership panels (exploratory; multiplicity discussed in Supplementary S4); separation/divorce outcomes; synthetic control on log respondent counts; entropy-balanced demographic reweighting; quarterly panels (pre 2018Q1– 2020Q4); summary index; max-*t* permutation correction across the outcome family; same-census-division pair restriction; restricted comparison pools; border states. Complete definitions and results in Supplementary S2–S4.

### Reversal test

The prediction, primary statistic and decision rule for the post-HB 4002 reversal test are specified in Supplementary S9 and deposited in a public registry (OSF), together with the frozen weight matrices, historical gap panels and analysis script, before the release of the 2025 BRFSS data (registration identifier in Supplementary S9).

### Data, code and reporting

All data sources are public. The complete replication package (Stata and Python, from raw downloads to figures) is available from the authors. The study follows STROBE guidance for observational studies where applicable (checklist: Supplementary S10). No ethics approval was required (de-identified public surveillance data).

## Data Availability

All data analyzed in this study are publicly available from the original sources: CDC Behavioral Risk Factor Surveillance System public-use files, CDC WONDER, CDC Vital Statistics Rapid Release, FBI NIBRS incident-level downloads, BLS Local Area Unemployment Statistics, CDC/NCHS Household Pulse Survey indicators, SAMHSA NSDUH state prevalence estimates, OxCGRT, and US Census/FRED. Replication code is available from the authors upon reasonable request. The pre-registered protocol for the post-repeal reversal test is deposited on OSF (registration osf.io/23cr8, under embargo until reporting).

https://www.cdc.gov/brfss/annual_data/annual_data.htm

https://wonder.cdc.gov/

https://www.cdc.gov/nchs/nvss/vsrr/drug-overdose-data.htm

https://cde.ucr.cjis.gov/LATEST/webapp/#/pages/downloads

https://www.bls.gov/lau/

https://www.samhsa.gov/data/

## Supplementary Information

### S1. Data and panel construction

We extract BRFSS public-use microdata (LLCP files) for 2011–2024. Table S1 lists variables and recodes. State-year cells aggregate all respondents with CDC final weights (_LLCPWT). The balanced panel retains 44 comparison units (43 states plus the District of Columbia) with complete outcome series over all 14 years, plus the two treated states (46 units in total); the five excluded states are Florida, Kentucky, New Jersey, Pennsylvania and Tennessee (incomplete series), and each treated state is evaluated within its own 45-unit panel that excludes the other. Item non-response on MENTHLTH is below 2% in every year and is dropped item-wise. External covariates: BLS LAUS unemployment; CDC VSRR synthetic-opioid (T40.4) death rates; CDC COVID mortality; Census/FRED population and income.

**Table S1.** Variables and definitions.

| Variable | BRFSS source | Definition |
| --- | --- | --- |
| FMD | MENTHLTH | 1[days $\geq 14$]; 88→0; 77, 99→missing |
| MHD | MENTHLTH | days of poor mental health, past 30 |
| Depression | ADDEPEV | ever told had a depressive disorder |
| Physical health | PHYSHLTH | days of poor physical health (same recodes) |
| Care renounced | MEDCOST | could not see a doctor because of cost, past 12m |
| Sex | SEXVAR/SEX1/SEX | harmonized across waves |
| Age bands | _AGEG5YR | 18–34, 35–64, 65+; 18–54 for household panels |
| Partnered | MARITAL | married or member of unmarried couple ($\in \{1, 6\}$) |
| Children | CHILDREN | children in household; 88→0; 99→missing |

Notes: Variable names as in the CDC BRFSS public-use codebooks; recodes applied identically in all survey years. FMD and MHD derive from the same `MENTHLTH` item. Source: CDC BRFSS annual public-use files, 2011–2024.

### S2. Complete estimation tables

Figure S1 shows the raw series behind the estimates: the frequent-mental-distress level of every panel unit, with the two treated states highlighted. Oregon’s climb through the distribution after 2021 is visible before any estimation. The figure also shows a rise common to most states from 2021, plausibly a pandemic aftermath; that common component is differenced out by construction, because the estimate is Oregon’s excess over states experiencing the same aftermath, and the differential version of the rival (a pandemic that hit Oregon harder) is tested directly by the stringency and school-closure adjustment in Supplementary S3. Table S2 reports the complete year-by-year gaps and placebo ranks: Oregon’s FMD gap is positive in each of the four post years, peaks in 2024 (+3.62 p.p.), and its post-average one-sided rank is 1/45, while the depression gaps stay flat or negative throughout. Table S3 shows the specification gates: Oregon remains between rank 1 and 4 of 45 under every pre-specified variant, and the joint pair test stays between *p* = 0.001 and 0.058. Table S4 collects the pooled and auxiliary inference: the pooled synthetic-DiD estimates are positive under both interval constructions, though the placebo-based intervals include zero, and the pair test survives the same-census-division restriction.

The depression outcome has the weakest pre-fit of the three (pair series above placebo bands in several pre years, driven by Washington); its post-2021 null is therefore quoted with that caveat wherever it appears. Detection floor for the diagnosis stock: with a lifetime-prevalence base of roughly 20% and annual new-diagnosis flows near 1–1.5% of adults, even a 20% rise in the flow moves the stock by under one percentage point over four years, close to the design’s granularity; the ADDEPEV null therefore excludes large flow changes only.

**Figure S1.**
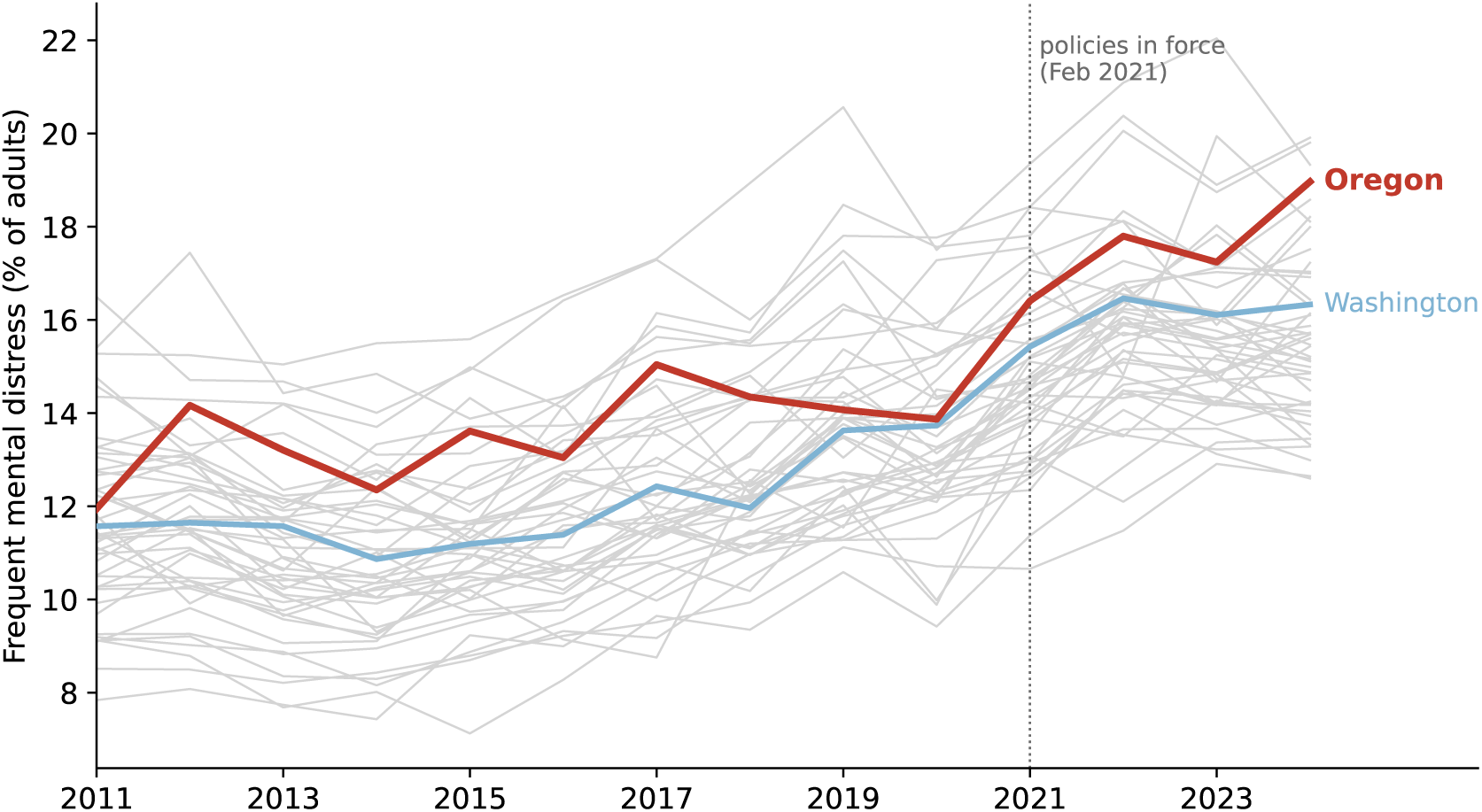
Frequent mental distress, raw levels, all 46 units, 2011–2024. Notes: Oregon (red) moves from the middle of the distribution toward its top after 2021; Washington (blue) rises less. Raw levels differ across states for compositional reasons, which is why the estimation compares each state with its own synthetic counterfactual rather than with the raw distribution. The 46 units are the 44 comparison units (grey) and the two treated states (colored). Weighted state-year shares. Source: authors’ calculations from CDC BRFSS public-use files.

**Table S2.**
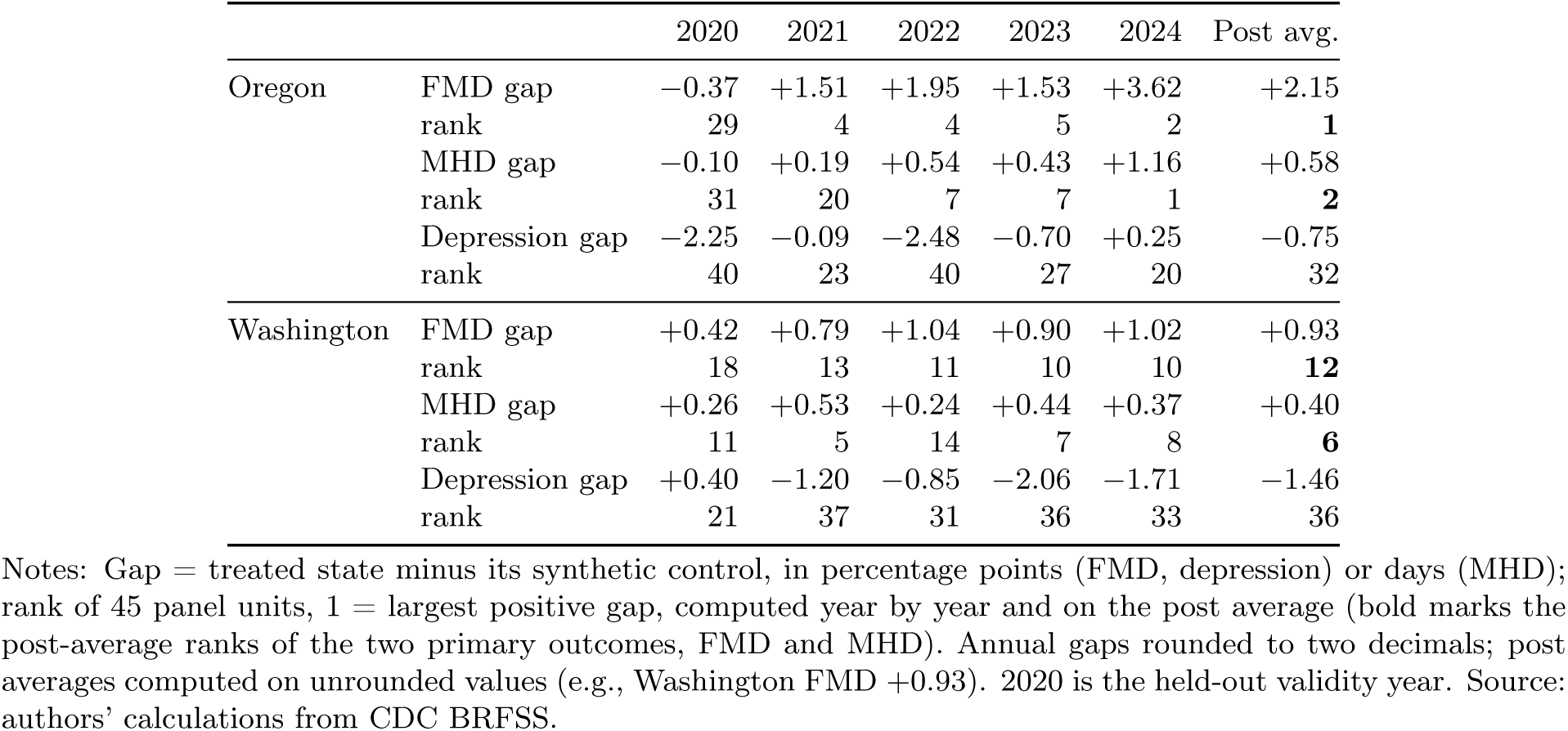
Year-by-year gaps and placebo ranks, all outcomes, both treated states.

**Table S3.**
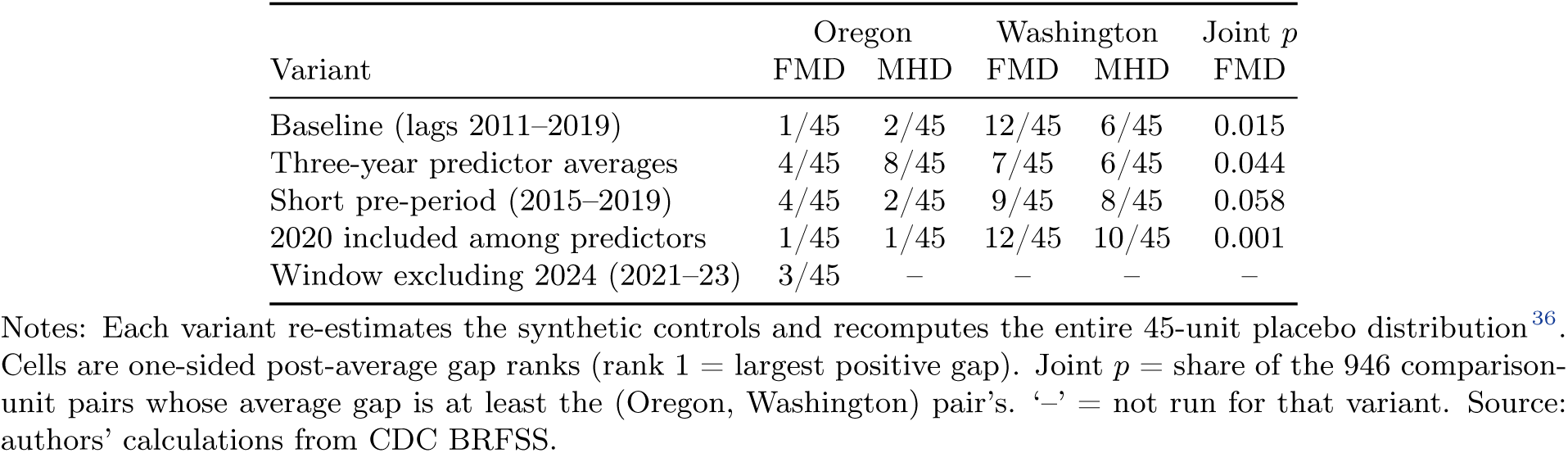
Specification gates.

**Table S4.** Pooled and auxiliary inference.

| Estimator / test | FMD | MHD |
| --- | --- | --- |
| Synthetic-DiD ATT (jackknife 95% CI) | +1.0 p.p. [+0.1, +1.9] | +0.30 d [+0.16, +0.44] |
| Synthetic-DiD ATT (placebo 95% CI) | [−0.3, +2.3] | [−0.04, +0.64] |
| Pair vs. 946 pairs, annual $p$ (2021–24) | 0.087 (82), 0.061 (58), 0.114 (108), 0.010 (9) | – |
| Pair pre-period deviation | 2013: $p$ = 0.039 (37/946; declared) | 2013: $p$ = 0.001 (1/946) |
| Summary index (mean $z$ of FMD, MHD) | rank 2/45 ($p$ = 0.044) | |
| Max-$t$ across outcome family | rank 3/45 ($p$ = 0.067) | |
| Same-division pairs only (112 pairs) | $p$ = 0.035 (4/113) | $p$ = 0.018 (2/113) |

Notes: Parenthetical counts are exact permutation counts, out of 946 donor pairs unless noted; same-division $p$ -values use the $(r+1)/(N+1)$ convention, counting the treated pair. Synthetic DiD = synthetic difference-in-differences ATT<sup>33</sup> with jackknife and placebo-based 95% intervals. Summary index = rank of the mean $z$ -score of FMD and MHD; max- $t$ = family-wise correction across the outcome family. Source: authors’ calculations from CDC BRFSS.

### S3. Inferential choices and diagnostics

Table S5 collects the inferential diagnostics and robustness estimates discussed in the main text and below: the headline result is robust to estimator changes (ridge-augmented rank 1/45 one-sided) and window choices, is weaker under the fit-normalized RMSPE-ratio statistic (11/43) and the covariates-as-predictors variant (3/45, with a markedly worse pre-fit), and the conformal procedure is uninformative in this short-post design.

Sub-state geography (SMART subsample). BRFSS public files carry no usable sub-state geography (the landline-only MSCODE variable is missing for *>*85% of respondents after 2017); the CDC SMART files provide metropolitan-area estimates with dedicated weights (_MMSAWT) for 2017–2019 and 2021–2024 (no 2020 release; 2020 is excluded from our design in any case). Among the 80 metropolitan areas with at least 150 respondents in all seven years, FMD changes from 2017–19 to 2021–24: Portland–Vancouver– Hillsboro +4.08 p.p. (rank 17/80; note the area includes Clark County, Washington, itself treated), Salem +4.65 p.p.. Oregon statewide rises +3.1 p.p. raw over the same window; with the Portland area roughly half of the state’s adult population, the implied remainder-of-state increase is about +2 p.p.. Series in smart_mmsa_fmd.csv.

Comparison-state weights for the baseline FMD synthetic controls. Weights below 0.001 are omitted from the display, which is why Washington’s listed weights sum to 0.993; full vectors are in the replication package. Oregon (pre-RMSPE 0.0059): Indiana 0.343, Mississippi 0.309, Delaware 0.162, South Carolina 0.103, New York 0.083. Washington (pre-RMSPE 0.0016): Arizona 0.232, California 0.224, Delaware 0.150, Iowa 0.147, Rhode Island 0.106, Louisiana 0.068, Idaho 0.066.

**Table S5.** Why gap-rank rather than RMSPE-ratio, and further diagnostics.

| Diagnostic | Result |
| --- | --- |
| Placebos with better pre-fit than Oregon (FMD) | 31 of 44 (pre-RMSPE OR 0.0059; placebo median 0.0043) |
| RMSPE-ratio test with $2\times$ fit filter | rank 11/43 ($p = 0.256$; the filter drops two ill-fitting units, hence 43) |
| In-time placebo (moving 4-year windows) | pre-windows ≥ post gap: 0 of 7 ($p = 0.125$, floor) |
| Restricted donor pool (pop. 0.5–3× OR, 26 states) | FMD rank 3/26; MHD rank 2/26 |
| Sampling (synth on log respondents) | rank 14/45 (central) |
| Entropy-balanced recomposition | reweighted increase ≥ raw increase |
| Quarterly panel (pre 2018Q1–2020Q4) | post rank 2/45 (FMD), 5/45 (MHD) |
| Two-sided rank on $\vert \hat{\tau}\vert $ (FMD) | 2/45 ($p = 0.044$) |
| Ridge-augmented SCM (LOOCV penalty) | gap +2.2 p.p.; rank 1/45 one-sided, 2/45 two-sided |
| Covariates as predictors (unemp., income, poverty + averaged lags) | gap +1.5 p.p.; rank 3/45 one-sided, 7/45 two-sided; pre-RMSPE 0.0084 |
| Physical-health days, full placebo loop | OR rank 21/45 ($p = 0.467$; 2020 rank 39/45) |
| Frequent physical distress ($\geq 14$ days) | OR rank 30/45 ($p = 0.667$) |
| Leave-one-year-out (FMD, drop 2021/22/23/24) | ranks 2, 2, 2, 3 of 45 |
| Joint pair test excluding 2024 (FMD) | $p = 0.027$ (26/946) |
| Residualized SCM (fentanyl+COVID+unemp partialled out) | OR rank 1/45 one-sided, 2/45 two-sided (gap +2.6 p.p.; 2020 rank 7/45) |
| Residualized SCM + OxCGRt stringency and school closing | OR rank 1/45 one-sided, 2/45 two-sided (gap +2.6 p.p.) |
| Conformal (CWZ)<sup>45</sup>: null-fit absorption diagnostic | post residuals +0.2/+1.1 p.p. (vs +1.6/+3.7 pre-only fit); pre-RMSPE degraded 0.61→0.78 p.p. (+28%; implementation-specific fit, cf. primary pre-RMSPE 0.59); $q/T = 4/13$, $J = 44 > T \Rightarrow$ test uninformative here |
| Quarterly 2024 split (FMD gap) | 2024Q1–Q2 (M110 only): +2.7 p.p.; 2023 avg.: +1.2 p.p. (annual-panel 2023: +1.5 p.p.; the quarterly panel matches on 2018–2020 and its gaps differ accordingly) |

Notes: One row per diagnostic; details in the surrounding text and Methods. Ranks are one-sided gap ranks of 45 units unless stated otherwise (43 under the $2\times$ pre-fit filter; 26 in the population-restricted pool; “two-sided” = rank of $|\text{gap}|$ ). $p$ = rank/number of units; exact pair counts in parentheses (x/946). Pre-RMSPE in outcome shares. Sources: authors’ calculations from CDC BRFSS; OxCGRt US state indices (stringency row).

### S4. The household channel (exploratory)

Table S6 reports the full subgroup battery: the increase concentrates among partnered women without children at home (+4.4 p.p., rank 3/45) and single men (rank 4/45), while mothers and partnered men are flat.

The two highest-ranked groups are partnered women and single men; partnered men and single women are unremarkable; among partnered women the gap is largest for those without children at home; and family composition itself does not move, so the pattern is not a divorce or selection artifact. Two formal tests now bracket the interpretation. (i) Effect modification: a microdata regression of FMD on Oregon*×*post interacted with the four sex-by-partnership groups (state and year effects by group, weights, SEs clustered by state) rejects homogeneity: omnibus *F* = 26.9, *p <* 0.001. Group-specific Oregon*×*post estimates from that regression: single men +1.95 p.p. (SE 0.18), partnered women +1.40 (net; interaction *−*0.55, SE 0.13), single women +0.42, partnered men +0.33: the same ordering as the synthetic-control battery. (ii)

Family-wise inference: taking each state’s maximum post-average gap across the eight household panels and ranking Oregon’s maximum (the childless-partnered-women gap, +4.4 p.p.) in that distribution gives rank 9/45, *p* = 0.20. Heterogeneity exists; no single subgroup is individually established. The pattern remains suggestive, and the subgroup battery for the reversal test is fixed in S9. Table S7 organizes the candidate mechanisms: ambient exposure to visible drug use retains the most consistent support, the household-exposure channel loses its direct tests (recorded IPV flat; no ACE differential), and a pure reporting artifact is bounded by the checks in Supplementary S8.

**Table S6.** Synthetic control by household structure.

| Panel | FMD gap | FMD rank | MHD gap | MHD rank |
| --- | --- | --- | --- | --- |
| *(a) Sex × partnership (all adults)* |  |  |  |  |
| Women, partnered | +1.7 p.p. | **3/45** | +0.57 | **3/45** |
| Women, not partnered | +1.6 p.p. | 12/45 | +0.17 | 17/45 |
| Men, partnered | +0.1 p.p. | 22/45 | +0.12 | 15/45 |
| Men, not partnered | +2.2 p.p. | **4/45** | +0.59 | **5/45** |
| *(b) Partnership × children, women 18–54* |  |  |  |  |
| Partnered, no children | +4.4 p.p. | **3/45** | +0.74 | 7/45 |
| Partnered, with children | +0.3 p.p. | 19/45 | −0.19 | 33/45 |
| Not partnered, with children | +1.3 p.p. | 18/45 | +0.32 | 16/45 |
| Not partnered, no children | +2.3 p.p. | 13/45 | +0.34 | 18/45 |
| *(c) Partnered women by age* |  |  |  |  |
| 25–54 | +1.6 p.p. | 8/45 | +0.36 | 11/45 |
| 55+ | +0.3 p.p. | 17/45 | +0.21 | 14/45 |
| *(d) Family-composition outcomes (women 25–54)* |  |  |  |  |
| Separated or divorced (share) | +0.4 p.p. | 19/45 |  |  |
| Separated only (share) | 0.0 p.p. | 27/45 |  |  |
| Partnered (share) | +0.6 p.p. | 10/45 |  |  |

Notes: Oregon post-average gap and one-sided rank of 45, from a separate synthetic control estimated on each subgroup panel identically to the main design. The battery is exploratory: no subgroup is individually significant after family-wise correction (text below); bold marks the highest ranks. Panels (b)–(d) restrict to the ages shown. Source: authors’ calculations from CDC BRFSS microdata.

**Table S7.**
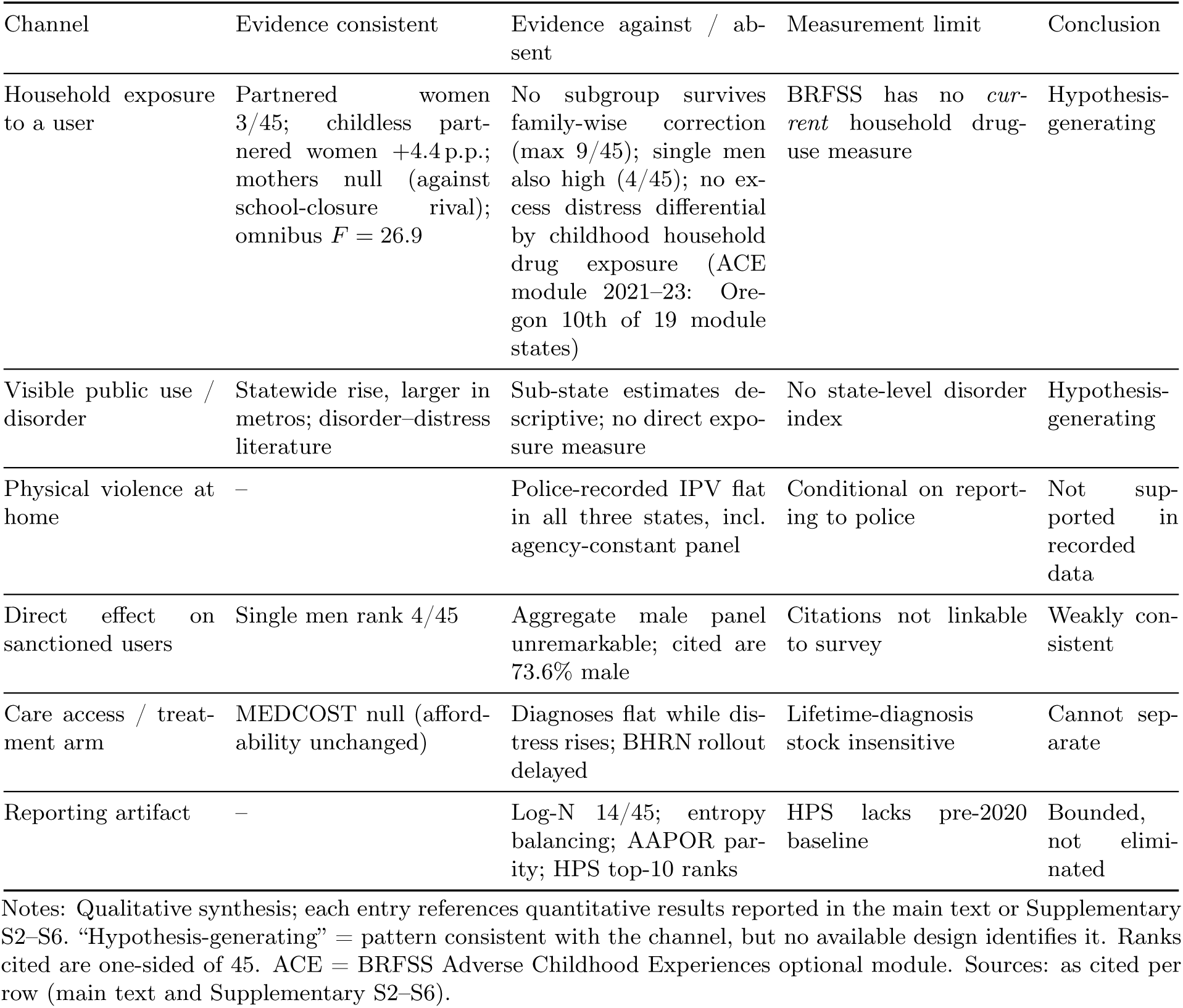
Candidate mechanisms: evidence for, evidence against, and what the available measures can capture.

### S5. Measured violence

IPV is constructed from FBI NIBRS incident-level bulk files (Oregon, Washington, Colorado, 2018–2024): female individual victims of aggravated or simple assault (offense codes 13A/13B) with victim–offender relationship spouse, common-law spouse, ex-spouse, boyfriend/girlfriend or ex-relationship. The parser exactly reproduces an independent earlier extraction for Oregon (month-by-month identity on all four series). Colorado serves as the untreated comparison: it shares the treated states’ legal-recreational-cannabis environment and a broadly comparable urban profile, never decriminalized other drugs, and is one of the few western states with complete incident-based NIBRS reporting throughout 2018–2024, so the series does not straddle the FBI’s 2021 retirement of summary reporting. Table S8 reports the monthly series: police-recorded IPV is flat through both treatments (Oregon +6.3% versus Colorado +7.2%), with no discontinuity at the treatment dates and no reversal after re-criminalization.

Comparison formula: the “vs. Colorado” column is the percentage-point difference of the two pre-to-post percentage changes. There is no discontinuity at the treatment dates: Washington’s full-*Blake* window (March–May 2021; ESB 5476 took effect 13 May 2021) averages 1,064 victims per month against the 1,141 pre mean, within normal monthly variation, and there is no reversal after re-criminalization. Measurement notes. (i) The FBI retired summary-based reporting nationally in January 2021, but Oregon, Washington and Colorado reported incident-based (NIBRS) data throughout 2018–2024, so the series do not straddle a reporting-system change. (ii) Agency composition: restricting to agencies (ORIs) with incidents in every year 2018–2024 retains 159 agencies in Oregon, 208 in Washington and 186 in Colorado, covering 96.8%, 99.1% and 98.5% of recorded female IPV victims respectively; on this constant panel the pre-to-post changes are +4.7% (OR), *−*1.7% (WA) and +6.8% (CO), confirming the full-panel conclusion (series in nibrs_*_ipv_constant.csv). (iii) Portland 911 domestic-violence dispatches double over the period (51 to 98–126 per month), but the rise begins in 2020 and does not reverse in late 2024; against flat statewide victim-level records, city-specific dispatch practice is the most plausible reading. Reporting caveat: if decriminalization reduced victims’ propensity to involve police, flat recorded IPV could mask a real increase.

**Table S8.**
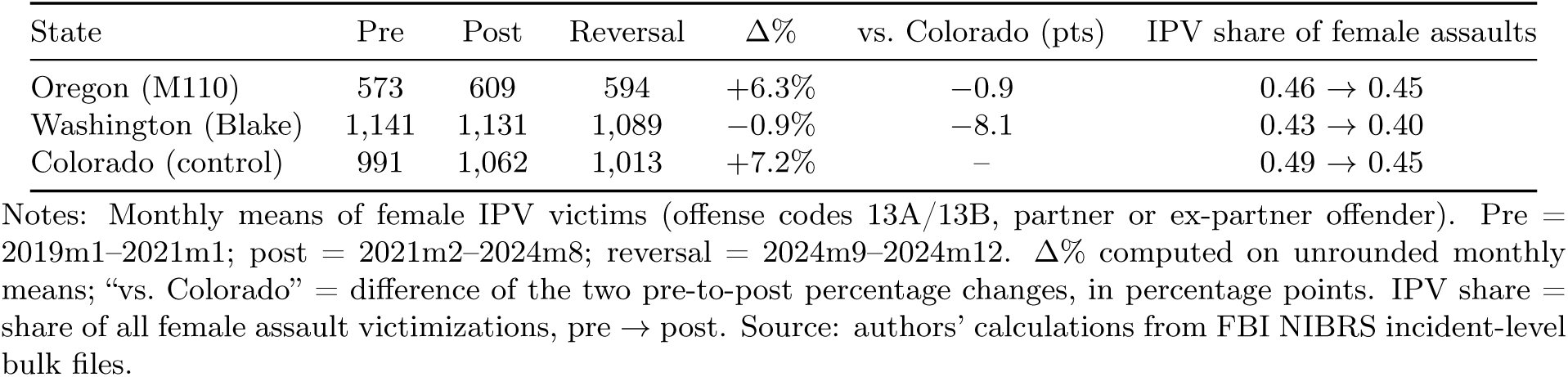
Female police-recorded IPV victims per month, NIBRS.

### S6. Suicide mortality

State-year suicide panel from CDC WONDER underlying-cause files (Injury Intent = Suicide): 1999–2020 (D76) and 2018–2024 (D158); the two sources are identical on the 2018–2020 overlap. The synthetic control design is identical to the main analysis (pre 2011–2019, 2020 excluded, post 2021–2024). Fifty-one jurisdictions (50 states and DC) enter; each treated state is evaluated within a 50-unit panel excluding the other treated state, and the joint test uses the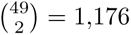 pairs of the 49 shared jurisdictions. Power note: the top five ranks of the 49 placebo post-average gaps begin at roughly +1.4 per 100,000; with Oregon’s baseline at 883 deaths in 2022 (crude rate 20.8 per 100,000; CDC age-adjusted rate 19.3), a policy effect below *∼*1.5 per 100,000 (*∼*7% of the baseline rate) would not reach the top ranks. The null excludes large effects, not all effects. Table S9 reports the synthetic-control results: Oregon’s post-average gap ranks 25/50, at the center of the placebo distribution, and the joint pair test is far from any conventional threshold.

Provisional 2025 data (extracted July 2026): Oregon 22.0 *→* 21.3 per 100,000, the ninth-largest decline of 51 jurisdictions; directionally consistent with reversal but uninformative given the overall null; we do not cite it as evidence in the main text.

**Table S9.**
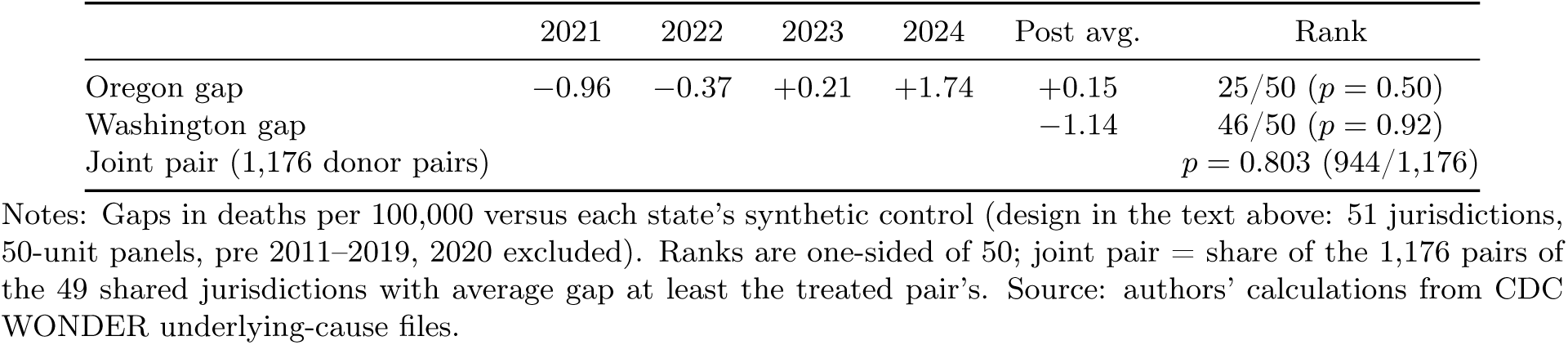
Suicide rates per 100,000: synthetic control results.

|  | 2021 | 2022 | 2023 | 2024 | Post avg. | Rank |
| --- | --- | --- | --- | --- | --- | --- |
| Oregon gap | –0.96 | –0.37 | +0.21 | +1.74 | +0.15 | 25/50 ($p = 0.50$) |
| Washington gap | | | | | –1.14 | 46/50 ($p = 0.92$) |
| Joint pair (1,176 donor pairs) | | | | | | $p = 0.803$ (944/1,176) |

Notes: Gaps in deaths per 100,000 versus each state’s synthetic control (design in the text above: 51 jurisdictions, 50-unit panels, pre 2011–2019, 2020 excluded). Ranks are one-sided of 50; joint pair = share of the 1,176 pairs of the 49 shared jurisdictions with average gap at least the treated pair’s. Source: authors’ calculations from CDC WONDER underlying-cause files.

### S7. Enforcement first stage

Sources: Oregon Judicial Department Measure 110 statistics ^4^ and Criminal Justice Commission reports ^39^. Class-E violation citations: approximately 12,400 circuit-court cases for offenses between February 2021 and August 2024 (*≈*3,400 per year; earlier OJD tallies report 4,451 by April 2023 and 6,177 by September 2023, i.e. *≈*2,000–2,400 per year over the first two and a half years). These replaced approximately 4,100 annual pre-2021 possession convictions (CJC). The large majority of citations resulted in failure to appear, and only a small fraction of recipients completed the health assessment (about 1% in early OJD tallies). Possession arrests fell by 68–83% depending on series and county. OJD demographic tabulations for cases with Class-E violations report 7,380 male (73.6%), 2,522 female (25.1%), 1 nonbinary and 125 missing-sex recipients (10,028 cases; this tabulation covers a shorter window than the *≈*12,400 cumulative circuit-court cases above, which extend through August 2024) ^4^: the sex contrast that anchors the paper’s comparison between the sanctioned population and the distressed population is taken directly from this official tabulation. Press accounts citing *∼*16,000 citations over three years include non-possession Class-E counts and duplicate filings; the circuit-court case series above is the consistent one. These magnitudes establish that the policy produced a large, discrete change in enforcement exposure, concentrated among men.

### S8. External triangulation and survey-quality diagnostics

The Household Pulse Survey (HPS) measures anxiety–depressive symptoms (PHQ-4 based, 7-day window) by state from April 2020, with an online panel and different weighting from BRFSS. Averaging published state estimates by calendar year, Oregon ranks 5th, 6th, 11th, 10th and 10th of 51 jurisdictions in 2020– 2024 (Washington: 16, 13, 24, 21, 18). Two readings follow. Against the reporting-artifact hypothesis: an independent instrument with a different mode also places Oregon near the top of the distress distribution throughout the post period. As a limit: HPS has no pre-2020 baseline, its 2020 values reflect the acute pandemic (and Oregon’s September 2020 wildfires), and its 2024 redesign breaks comparability, so it cannot corroborate the pre–post *change* that the BRFSS design identifies. Source: CDC/NCHS Indicators of Anxiety or Depression, data.cdc.gov (series 8pt5-q6wp).

#### NSDUH triangulation

The National Survey on Drug Use and Health (NSDUH) measures substance use and mental illness within the same instrument, with state estimates published in two-year waves. Table S10 reports Oregon’s within-wave national rank: persistently at the top of the distribution of non-marijuana illicit use, and moving to the top of the mental-illness distribution between the 2021–22 and 2022–23 waves, a within-redesign comparison.

#### Response-rate diagnostics

Weighted AAPOR combined (landline + cell) response rates (RR4) from the CDC BRFSS response-rate tables, published at state level from 2015; Table S11 reports them: Oregon’s post-2021 deviations from the national median lie well within its pre-treatment range. Post-treatment, both treated states track the national median: Oregon’s deviations in 2021–2024 (*−*0.7, +1.9, *−*6.4, +0.1 points) lie well within its pre-treatment range (*−*10.1 to +9.6), and Washington’s post rates are within one point of the median in every year. There is no systematic post-2021 response-rate divergence that could generate differential selection into the survey; this complements the log-respondents synthetic control (rank 14/45) and the entropy-balancing recomposition test.

**Table S10.** Oregon’s national rank in the NSDUH state estimates.

|  | 2016–17 | 2018–19 | 2021–22 | 2022–23 |
| --- | --- | --- | --- | --- |
| Illicit drug use other than marijuana (past month) | 3 | 3 | 2 | 2 |
| Any mental illness | 2 | 3 | 3 | 1 |
| Serious mental illness | 7 | 9 | 6 | 2 |
| Substance use disorder | 4 | 6 | 6 | 4 |

Notes: Adults 18+; rank 1 = highest prevalence among 51 jurisdictions (50 states and DC). Ranks are within-wave orderings of point estimates, not significance tests; the 2021 multimode redesign makes levels non-comparable across the break (the 2021–22 and 2022–23 waves are mutually comparable). Source: SAMHSA, NSDUH State Prevalence Estimates, releases 2016–2017 through 2022–2023, samhsa.gov/data.

**Table S11.** BRFSS weighted AAPOR combined response rates (RR4, %), treated states vs. national median.

|  | 2015 | 2016 | 2017 | 2018 | 2019 | 2020 | 2021 | 2022 | 2023 | 2024 |
| --- | --- | --- | --- | --- | --- | --- | --- | --- | --- | --- |
| Oregon | 50.0 | 56.6 | 45.9 | 39.8 | 46.5 | 50.1 | 43.3 | 47.0 | 38.2 | 43.9 |
| Washington | 36.0 | 39.3 | 39.6 | 50.0 | 47.1 | 47.6 | 44.0 | 45.0 | 45.0 | 44.6 |
| Median (all states) | 47.2 | 47.0 | 45.8 | 49.9 | 49.4 | 47.9 | 44.0 | 45.1 | 44.6 | 43.8 |
| Oregon – median | +2.8 | +9.6 | +0.1 | –10.1 | –2.9 | +2.2 | –0.7 | +1.9 | –6.4 | +0.1 |

Notes: Weighted AAPOR combined (landline + cell) response rates, RR4, in percent; median across all participating states. Values transcribed from the annual CDC BRFSS summary data-quality reports (the 2017 median is computed from the published state values; all other medians as published). Source: CDC BRFSS data-quality reports, 2015–2024.

### S9. Pre-specification of the HB 4002 reversal test

The full protocol (version 2.2, 7 August 2026) is deposited on OSF together with the frozen weight matrices and the analysis script (OSF registration osf.io/23cr8, deposited 8 August 2026 under embargo until August 2028), before the release of the 2025 BRFSS data (expected August–September 2026). Summary, verbatim on the decision categories. *Prediction*: under the policy-effect hypothesis, Oregon’s FMD gap closes once the re-criminalized regime covers the full survey year (2025; 2024 contains four re-criminalized months). *Primary statistic*: the level rank *R*_25_ of Oregon’s 2025 single-year gap among the 45 units (2021–2024 level ranks: 4, 4, 5, 2), chosen because placebo gaps are largely transitory (pre-period mean-reversion *β ≈ −*0.86; a change measured against 2024, Oregon’s largest year, would register a decline nearly automatically and is reported descriptively only), while Oregon’s four-year persistence contradicts the transience model. *Co-primary*: Δ*^′^* = gap(2025) *−* mean gap(2021–24), ranked among the 45 analogues, with the pre-period calibration frozen in the protocol. *Decision rule* (FMD only; five verbatim categories, evaluated in fixed order, first match wins): evidence against the mechanism’s reversibility (*R*_25_ *≤* 5 with gap at or above the 2021–24 average); supportive of reversal (*R*_25_ *≥* 11, decline *≥* 0.72 p.p., Δ*^′^* rank *≤* 10*/*45, residualized co-verdict agrees); reversal not separable from drug-supply improvement (as supportive, but the residualized co-verdict disagrees); weakly consistent with reversal (floor met, with *R*_25_ 6–10, or *R*_25_ *≥* 11 with unremarkable Δ*^′^* rank, or *R*_25_ *≤* 5 with Δ*^′^* rank *≤* 3*/*45); no evidence of reversal (all remaining configurations). *Qualifiers*: residualized co-verdict computed from machinery frozen at deposit (coefficients, weights, historical gaps); Washington regional check (its July 2023 re-criminalization makes a Washington decline ambiguous between regional confounding and the mechanism, so the check adds the fixed qualifier “not regionally distinguishable” without altering the category); interview-month timing test around the 1 September 2024 effective date. *Asymmetry note*: the test is asymmetric by design. A decline timed to re-criminalization would strengthen the policy reading, though coincident improvements in the drug-supply environment could contribute to it; a null reversal weakens the mechanism but admits an alternative reading, because a policy environment that raises distress need not lower it symmetrically on repeal (hysteresis, consistent with the Washington 2023 experience). The decision rule therefore treats a realized reversal as supportive and a non-reversal as informative but not decisive. *Hierarchy*: MHD analogues and two-sided versions are reported but cannot change the verdict; the exploratory subgroup battery is fixed (women; men; 18–34; partnered women without children at home), evaluated with a max-statistic; no other subgroup will be reported.

### S10. Reporting checklist

The completed STROBE checklist is included in the replication package.

